# Disparities in Access to, Use of, and Quality of Rehabilitation Care Following Stroke: A Scoping Review

**DOI:** 10.1101/2023.10.31.23297883

**Authors:** Janet K. Freburger, Elizabeth R. Mormer, Kristin Ressel, Shuqi Zhang, Anna M. Johnson, Amy M. Pastva, Rose L. Turner, Peter C. Coyle, Cheryl D. Bushnell, Pamela W. Duncan, Sara B. Jones Berkeley

## Abstract

Despite the clear utility of stroke rehabilitation, our understanding of the extent of disparities in rehabilitation care across specific sociodemographic characteristics (e.g., by age, sex, race, ethnicity, rurality, socioeconomic status) is limited. In this review, we aimed to synthesize the current evidence on disparities in referral to, use of, and quality of rehabilitation following stroke; identify research gaps in our understanding of disparities in rehabilitation care; and make recommendations for future research to advance the field in identifying, understanding the causes of, and minimizing disparities. We searched three electronic databases from 1997 to 2023 for cohort, case-control, cross-sectional, observational, and mixed-method studies that examined disparities in rehabilitation care following stroke. From 7,853 records screened, we identified 49 studies that met our inclusion criteria. Findings from the studies we reviewed varied, with the most consistent findings indicating that individuals with lower socioeconomic status and those living in rural areas (vs urban) are less likely to receive rehabilitation care following stroke. Findings on racial, ethnic, and sex disparities varied with some consistent findings among specific subgroups (e.g., studies examining care in the outpatient setting) and when examining results from analyses that controlled for functional status and other measures of “need” for therapy. Several gaps in our understanding of disparities in rehabilitation care following stroke were identified. Recommendations for future work were also provided.

## INTRODUCTION

There are more than 795,000 new or recurrent strokes each year in the United States (US), and stroke is a major cause of long-term disability in adults.^1–3^ Approximately 80-90% of adult stroke survivors have residual mobility impairments, ^4,5^ and approximately one-third have cognitive impairments.^6^ Post-stroke impairments in mobility and cognition affect quality of life, interfere with activities of daily living, and reduce social and occupational participation.^7^ Residual impairments also increase the risk of inactivity, falls, and hospital readmission.^8–11^

Physical therapists (PT), occupational therapists (OT), and speech language pathologists (SLP) play instrumental roles in maximizing patients’ functional status and addressing needs following stroke. Together with patients, they set goals and design and implement interventions to optimize mobility, cognitive function, and social and occupational participation. They also provide education on physical activity, fall prevention, and caregiving for stroke survivors.^5^

Evidence supporting the efficacy and effectiveness of rehabilitation following stroke to improve function and prevent adverse outcomes (e.g., falls, deconditioning, pneumonia, hospital readmissions) is quite robust, with several systematic reviews, randomized controlled trials, and observational studies reported in the literature.^12–18^ Clinical practice guidelines from the American Heart Association/American Stroke Association^18^ and the US Department of Veteran’s Affairs (VA)/Department of Defense ^19^ underscore the value of rehabilitation following stroke, and assessment for or receipt of stroke rehabilitation is one of the eight quality metrics evaluated by the Joint Commission for the certification of comprehensive stroke centers.^20^

Despite the clear utility of stroke rehabilitation, our understanding of the extent of disparities in rehabilitation care across specific sociodemographic characteristics (e.g., by age, sex, race, ethnicity, rurality, socioeconomic status [SES]) is limited. While several reviews have been conducted on disparities in acute stroke management and stroke prevention,^21–26^ to our knowledge, none exists for disparities in stroke rehabilitation. Filling this gap is particularly important given that the incidence of stroke and outcomes following stroke disproportionally affect certain demographic and socioeconomic groups identified as disparity populations.^26,27^ ^28,29^ Furthermore, advances in post-stroke care and secondary prevention have decreased mortality rates for stroke,^30^ leading to an increased prevalence of stroke survivors, many of whom have functional limitations.

A considerable challenge in studying disparities in stroke rehabilitation is that rehabilitation is complex, being delivered in multiple settings, by multiple provider types, and across time. (Figure 1) The quality of care may also vary. Disparities in rehabilitation care, therefore, can be assessed in different settings, at different time points during recovery, and from the perspectives of one or more provider types. Disparities in referral to and utilization of rehabilitation care can also be examined in different ways, from the simple measure of use or receipt of care to more nuanced measures of the frequency, intensity, duration, or type of care. Disparities in the time to initiation of care relative to stroke onset and/or transitions across settings may exist as well. Finally, one can also examine disparities in the quality of care delivered, in the context of the patient’s needs (e.g content, frequency, and duration of care).

**Figure 1.**
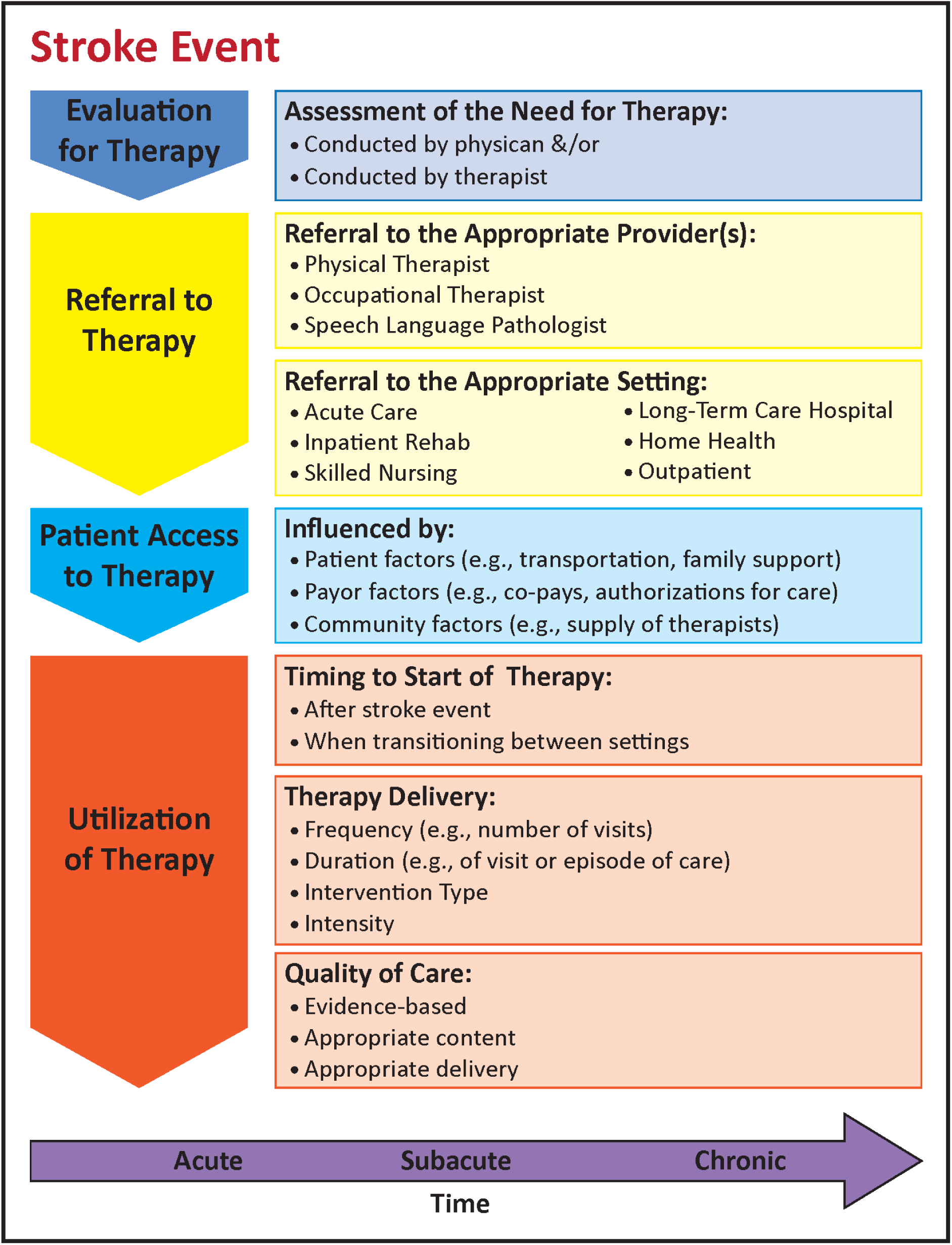
Rehabilitation Disparities Framework Conceptual Model. This figure is a conceptual model illustrating the process of rehabilitation care following a stroke event. The arrow indicates that these processes occur across time in the acute, subacute, and chronic time periods.

Using our conceptual model for rehabilitation care following stroke as a framework (**Figure 1**), we conducted a scoping review to: a) synthesize the current evidence on disparities in referral to, use of, and quality of rehabilitation following stroke; b) identify research gaps in our understanding of disparities in rehabilitation care, and c) make recommendations for future research to advance the field in identifying, understanding the causes of, and minimizing disparities.

## METHODS

We conducted a scoping review of disparities in rehabilitation care and reported items according to the PRISMA Extension for Scoping Review guidelines.^31^ Our protocol was not registered. We considered the following disparity populations based on definitions from the National Institutes of Health (NIH)^28^ and the Agency for Healthcare Research and Quality: racial minorities; ethnic minorities; sex and gender minorities; older population; low income or socioeconomically disadvantaged populations; and geographic minorities (inner city/rural).^29^ Because a significant consequence of stroke is disability, we did not include disabled individuals a disparity population.

Rehabilitation care was defined as care delivered by a PT, OT and/or SLP in inpatient, outpatient, or home settings; care referred to as rehabilitation, with no other information on provider type; or post-acute care received in an inpatient rehabilitation facility (IRF), skilled nursing facility (SNF), long-term care hospital (LTH), or the home via home health (HH).

### Information Sources and Search

Detailed searches of MEDLINE (Ovid), Embase (Elsevier), and Cumulative Index to Nursing and Allied Health Literature (CINAHL, EBSCO) were conducted by an experienced health sciences librarian. Because healthcare delivery is influenced by healthcare policy, which varies by country and over time, we limited our review to English only publications of studies conducted in the US from January 1, 1997 to March 15, 2023. We began the review in 1997 because that would cover a roughly 25-year period and, in 1997, the Balanced Budget Act led to significant changes in Medicare policies for rehabilitation care delivery in the US.^29^

Cohort, case-control, cross-sectional, observational, and mixed-method studies were eligible for inclusion; abstracts, reviews, editorials, commentaries, protocols, and non-research letters were excluded. Systematic and scoping reviews were included for background information but were not eligible for screening. We limited studies to those with adults (≥18 years) with stroke.

A combination of Medical Subject Headings (MeSH) terms and title, abstract, and keywords were used to develop the initial Medline search, which was validated against a known set of relevant studies. The Medline search was then adapted to the other databases. We used a validated geographic search hedge to remove non-US studies^32^ and added sensitive search hedges to remove pediatric studies and animal studies (Embase only). Primary strings included in our search were “stroke”; “health care disparities”; “rehabilitation”; and strings for our disparity populations (e.g., “race”, “ethnicity”). We adapted a “stroke” search string from Clark et al.^33^ and a Hispanic/Latinx search string from the MLA Latinx Caucus.^34^ We used a polyglot search translator to assist in search translations.^35^ **Supplemental Tables S1-S3** outline the full search strategy for each database.

### Selection of Sources of Evidence

Database records were exported to an EndNote library, ^36^ and any duplicate records were removed using Bramer^37^ and Amsterdam Efficient Deduplication methods.^38^ Remaining records were uploaded to DistillerSR.^39^ The first-level screening of studies was performed by a subset of study author in pairs (JF, AJ, AP, KR, SJB, EM) who reviewed titles and abstracts using a screening form. The screening process and form were piloted on 50 references and revised until interrater reliability among the screeners was >0.60 using Cohen’s kappa statistic.

Studies were excluded if they were conducted outside of the US, were not limited to participants with stroke (e.g., participants with both stroke and cardiovascular disease), were conducted on individuals <18 years of age or did not examine disparities in rehabilitation care. Screeners retained articles if there was uncertainty about whether exclusion criteria were met. Disagreements were resolved within screening pairs and brought to the larger group if consensus was not reached. If consensus was not able to be reached by the larger group, the article was retained for Level 2 screening.

Level 2 screening, which consisted of full-text review, was completed by four authors (AJ, EM, KR, SZ). References from Level 2 that were unclear were adjudicated by senior authors JF and AP to determine final inclusion for data charting.

### Data Charting and Synthesis

Retained articles were charted using DistillerSR^39^. We developed a data charting form and instruction manual using selected manuscripts for guidance (**Supplemental Table S4)**. The form was piloted by seven study authors who provided feedback, and revisions were made. Each study was charted by one of four study authors (AP, EM, PC, SZ). Data were primarily synthesized quantitatively using descriptive statistics.

## RESULTS

Our search strategy yielded 7583 articles after duplicates were removed. These were screened, and 473 were retained for full-text review. Forty-nine articles published between 1997 and 2023 met inclusion criteria and were included in our review (Figure 2). Most (n=30, 61.2%) were published in 2010 or later. Less than half (n=23, 46.9%) had stated objectives to examine disparities in rehabilitation care, and 37 (74.5%) had stated objectives to examine rehabilitation use. A complete list of the authors, title, publication year, and objectives are presented in **Supplemental Table S5**.

**Figure 2.**
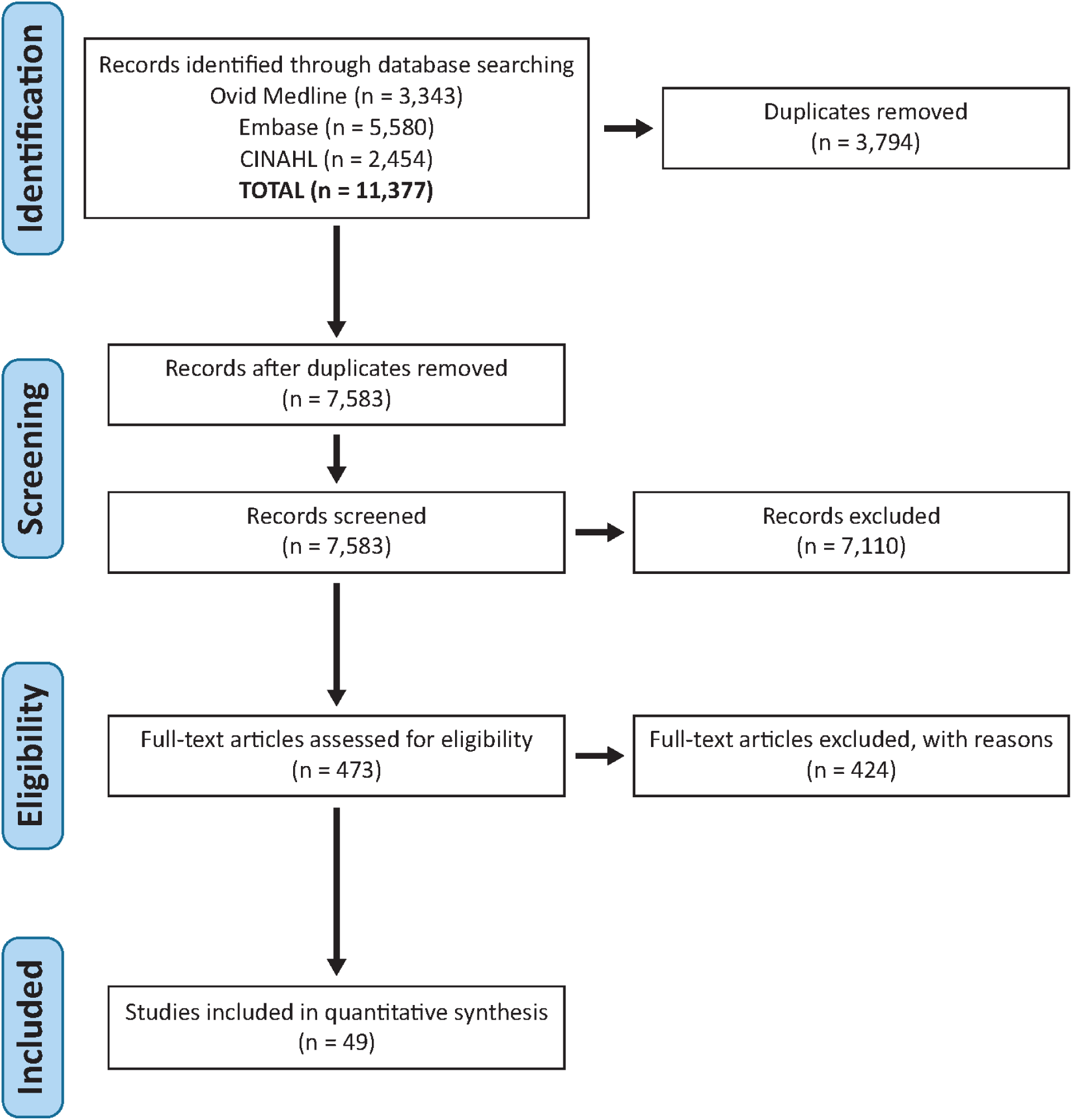
PRISMA Flow Diagram. This diagram provides a visual representation of the study selection process, following the PRISMA (Preferred Reporting Items for Systematic Reviews and Meta-Analyses) guidelines. The flow chart traces the number of records identified, screened, included, and excluded at various stages of the review, offering clarity on the filtration process leading to the final set of studies considered.

### Objectives, Study Design, and Data Source

**Table 1** summarizes the characteristics of studies overall and stratified by type(s) of disparities examined. Disparities by race and/or ethnicity were the most common types examined, (n=43, 87.8%); urban/rural disparities were the least common (n=8, 16.3%). For study design, 22 (44.9%) used prospective cohort, 14 (28.6%) used cross-sectional, and 13 (26.5%) used retrospective cohort design. The distribution of study designs varied by the type of disparity examined. Notably, studies examining urban/rural disparities did not include any prospective cohort studies.

**Table 1.**
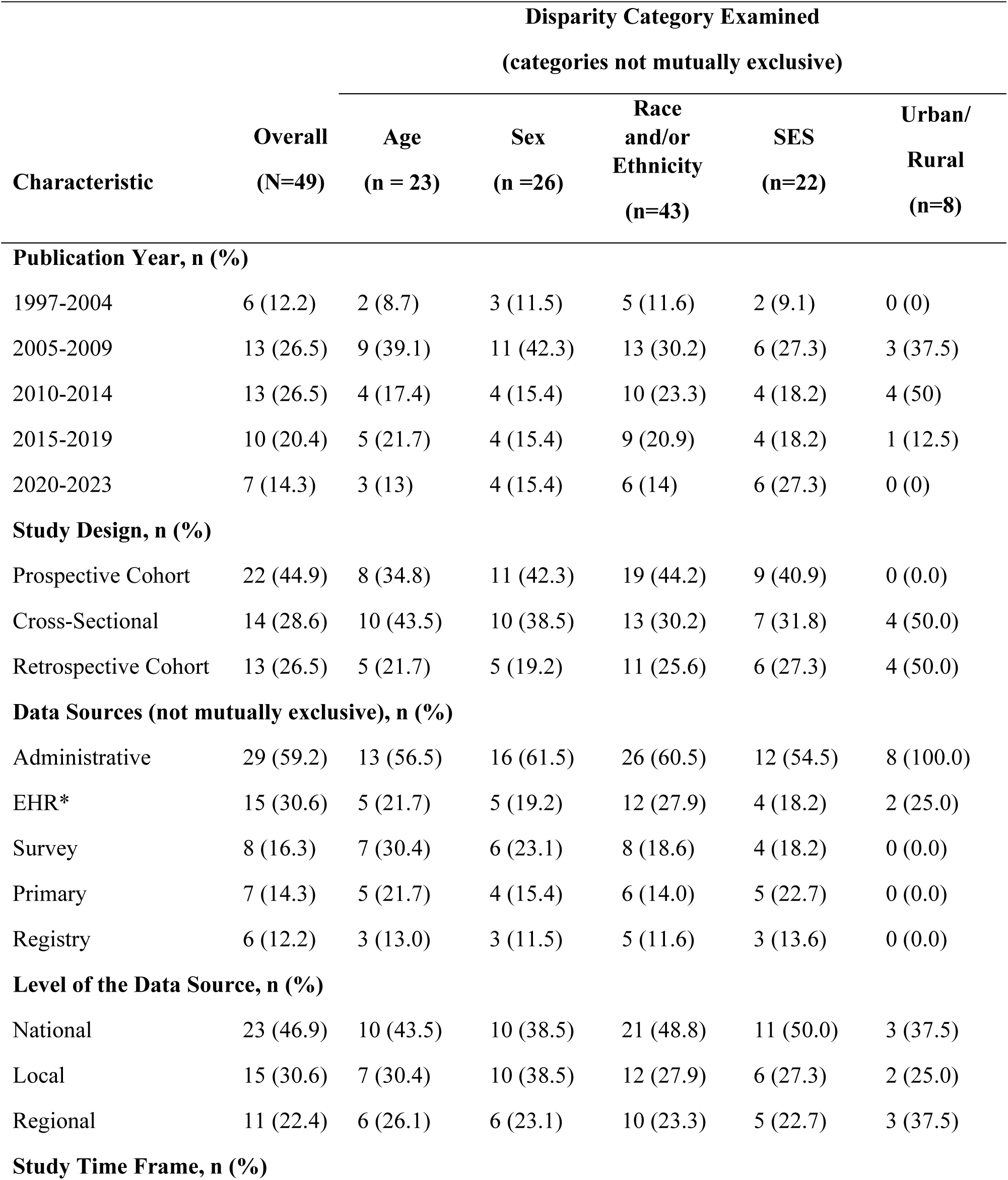
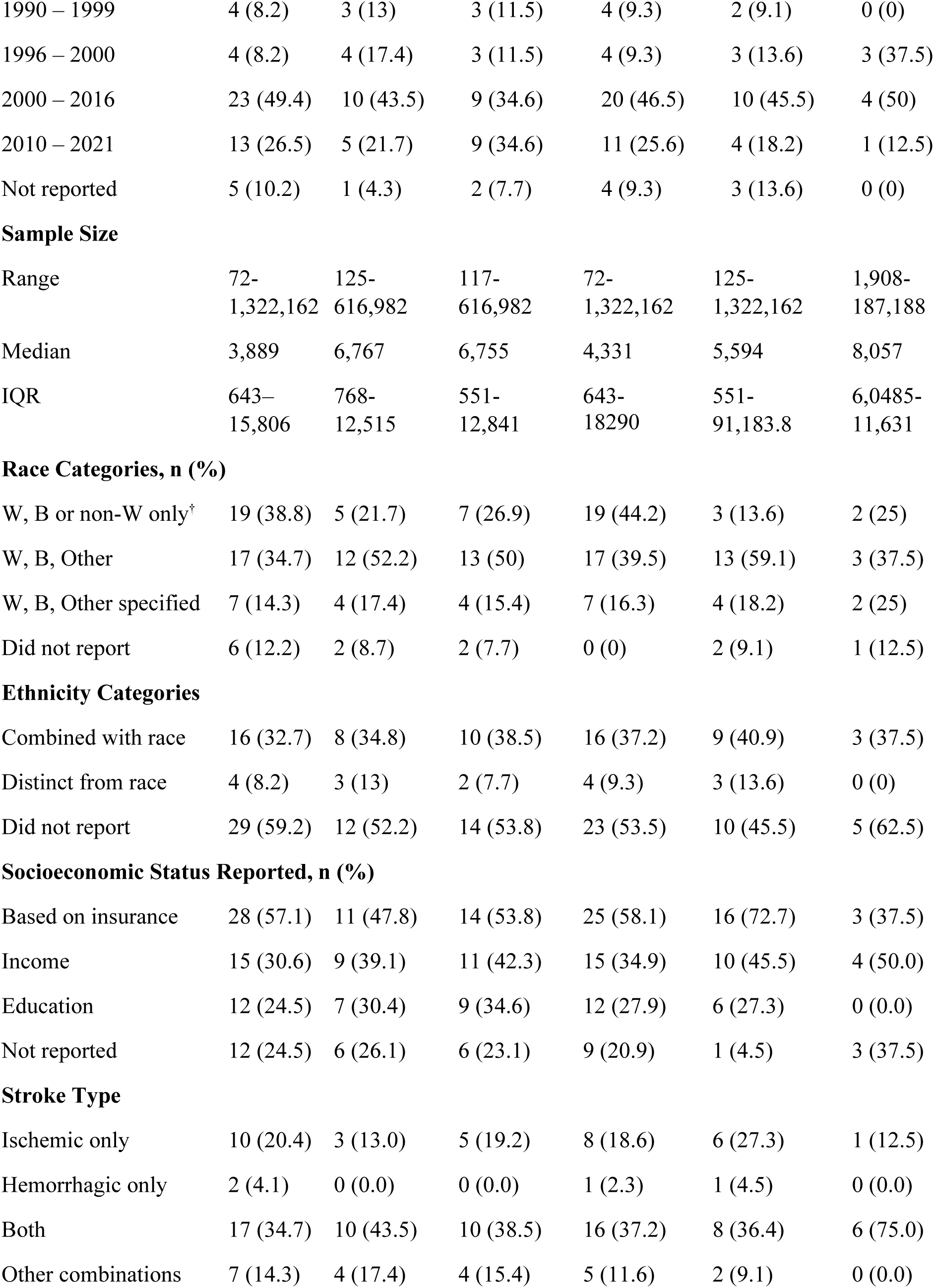
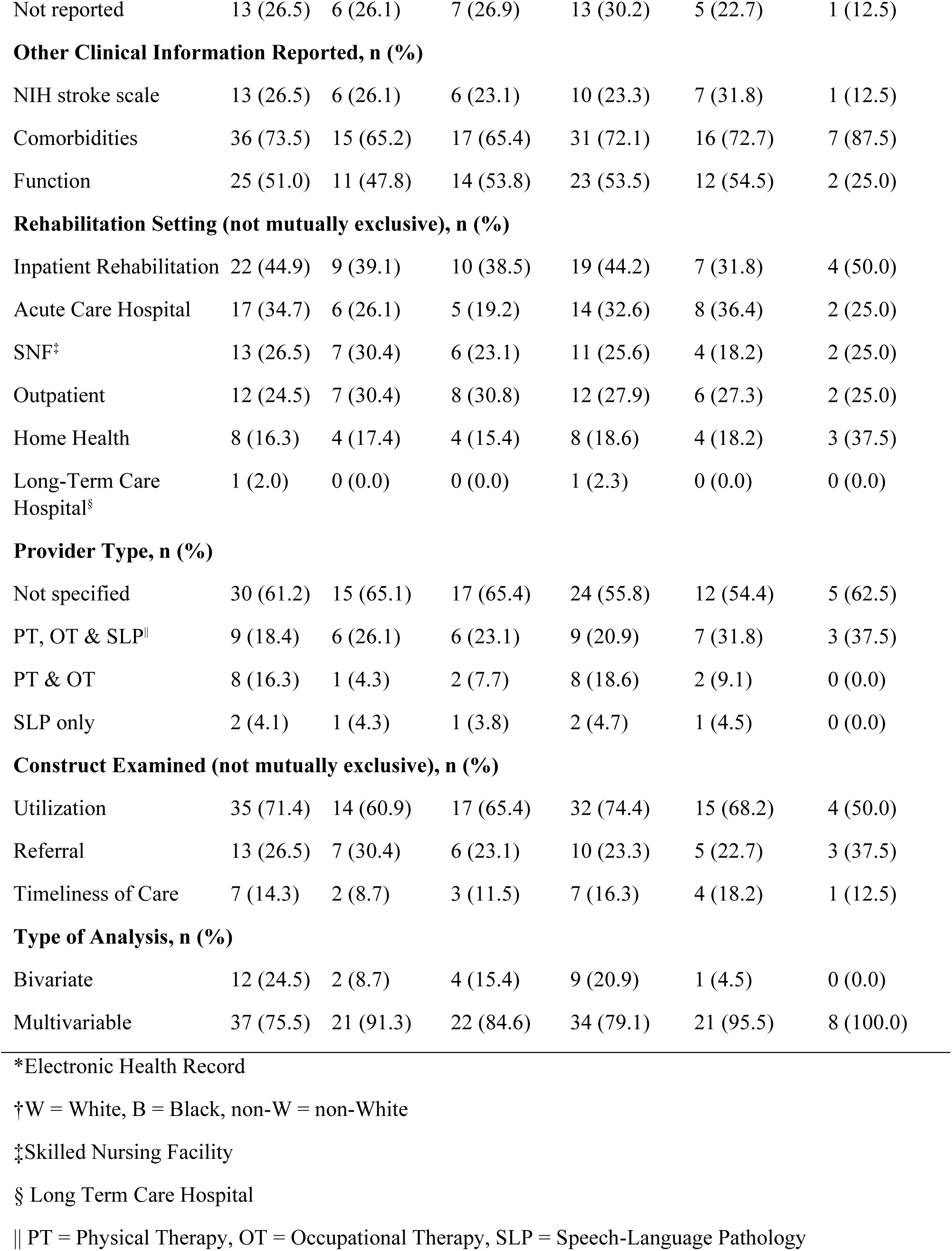
Study Characteristics.

| Characteristic | Disparity Category Examined<br>(categories not mutually exclusive) |  |  |  |  |  |
| --- | --- | --- | --- | --- | --- | --- |
|  | Overall<br>(N=49) | Age<br>(n = 23) | Sex<br>(n =26) | Race<br>and/or<br>Ethnicity<br>(n=43) | SES<br>(n=22) | Urban/<br>Rural<br>(n=8) |
| **Publication Year, n (%)** |  |  |  |  |  |  |
| 1997-2004 | 6 (12.2) | 2 (8.7) | 3 (11.5) | 5 (11.6) | 2 (9.1) | 0 (0) |
| 2005-2009 | 13 (26.5) | 9 (39.1) | 11 (42.3) | 13 (30.2) | 6 (27.3) | 3 (37.5) |
| 2010-2014 | 13 (26.5) | 4 (17.4) | 4 (15.4) | 10 (23.3) | 4 (18.2) | 4 (50) |
| 2015-2019 | 10 (20.4) | 5 (21.7) | 4 (15.4) | 9 (20.9) | 4 (18.2) | 1 (12.5) |
| 2020-2023 | 7 (14.3) | 3 (13) | 4 (15.4) | 6 (14) | 6 (27.3) | 0 (0) |
| **Study Design, n (%)** |  |  |  |  |  |  |
| Prospective Cohort | 22 (44.9) | 8 (34.8) | 11 (42.3) | 19 (44.2) | 9 (40.9) | 0 (0.0) |
| Cross-Sectional | 14 (28.6) | 10 (43.5) | 10 (38.5) | 13 (30.2) | 7 (31.8) | 4 (50.0) |
| Retrospective Cohort | 13 (26.5) | 5 (21.7) | 5 (19.2) | 11 (25.6) | 6 (27.3) | 4 (50.0) |
| **Data Sources (not mutually exclusive), n (%)** |  |  |  |  |  |  |
| Administrative | 29 (59.2) | 13 (56.5) | 16 (61.5) | 26 (60.5) | 12 (54.5) | 8 (100.0) |
| EHR* | 15 (30.6) | 5 (21.7) | 5 (19.2) | 12 (27.9) | 4 (18.2) | 2 (25.0) |
| Survey | 8 (16.3) | 7 (30.4) | 6 (23.1) | 8 (18.6) | 4 (18.2) | 0 (0.0) |
| Primary | 7 (14.3) | 5 (21.7) | 4 (15.4) | 6 (14.0) | 5 (22.7) | 0 (0.0) |
| Registry | 6 (12.2) | 3 (13.0) | 3 (11.5) | 5 (11.6) | 3 (13.6) | 0 (0.0) |
| **Level of the Data Source, n (%)** |  |  |  |  |  |  |
| National | 23 (46.9) | 10 (43.5) | 10 (38.5) | 21 (48.8) | 11 (50.0) | 3 (37.5) |
| Local | 15 (30.6) | 7 (30.4) | 10 (38.5) | 12 (27.9) | 6 (27.3) | 2 (25.0) |
| Regional | 11 (22.4) | 6 (26.1) | 6 (23.1) | 10 (23.3) | 5 (22.7) | 3 (37.5) |
| **Study Time Frame, n (%)** |  |  |  |  |  |  |
| 1990 – 1999 | 4 (8.2) | 3 (13) | 3 (11.5) | 4 (9.3) | 2 (9.1) | 0 (0) |
| 1996 – 2000 | 4 (8.2) | 4 (17.4) | 3 (11.5) | 4 (9.3) | 3 (13.6) | 3 (37.5) |
| 2000 – 2016 | 23 (49.4) | 10 (43.5) | 9 (34.6) | 20 (46.5) | 10 (45.5) | 4 (50) |
| 2010 – 2021 | 13 (26.5) | 5 (21.7) | 9 (34.6) | 11 (25.6) | 4 (18.2) | 1 (12.5) |
| Not reported | 5 (10.2) | 1 (4.3) | 2 (7.7) | 4 (9.3) | 3 (13.6) | 0 (0) |
| Sample Size |  |  |  |  |  |  |
| Range | 72-1,322,162 | 125-616,982 | 117-616,982 | 72-1,322,162 | 125-1,322,162 | 1,908-187,188 |
| Median | 3,889 | 6,767 | 6,755 | 4,331 | 5,594 | 8,057 |
| IQR | 643–<br>15,806 | 768-12,515 | 551-12,841 | 643-18290 | 551-91,183.8 | 6,0485-11,631 |
| Race Categories, n (%) |  |  |  |  |  |  |
| W, B or non-W only<sup>†</sup> | 19 (38.8) | 5 (21.7) | 7 (26.9) | 19 (44.2) | 3 (13.6) | 2 (25) |
| W, B, Other | 17 (34.7) | 12 (52.2) | 13 (50) | 17 (39.5) | 13 (59.1) | 3 (37.5) |
| W, B, Other specified | 7 (14.3) | 4 (17.4) | 4 (15.4) | 7 (16.3) | 4 (18.2) | 2 (25) |
| Did not report | 6 (12.2) | 2 (8.7) | 2 (7.7) | 0 (0) | 2 (9.1) | 1 (12.5) |
| Ethnicity Categories |  |  |  |  |  |  |
| Combined with race | 16 (32.7) | 8 (34.8) | 10 (38.5) | 16 (37.2) | 9 (40.9) | 3 (37.5) |
| Distinct from race | 4 (8.2) | 3 (13) | 2 (7.7) | 4 (9.3) | 3 (13.6) | 0 (0) |
| Did not report | 29 (59.2) | 12 (52.2) | 14 (53.8) | 23 (53.5) | 10 (45.5) | 5 (62.5) |
| Socioeconomic Status Reported, n (%) |  |  |  |  |  |  |
| Based on insurance | 28 (57.1) | 11 (47.8) | 14 (53.8) | 25 (58.1) | 16 (72.7) | 3 (37.5) |
| Income | 15 (30.6) | 9 (39.1) | 11 (42.3) | 15 (34.9) | 10 (45.5) | 4 (50.0) |
| Education | 12 (24.5) | 7 (30.4) | 9 (34.6) | 12 (27.9) | 6 (27.3) | 0 (0.0) |
| Not reported | 12 (24.5) | 6 (26.1) | 6 (23.1) | 9 (20.9) | 1 (4.5) | 3 (37.5) |
| Stroke Type |  |  |  |  |  |  |
| Ischemic only | 10 (20.4) | 3 (13.0) | 5 (19.2) | 8 (18.6) | 6 (27.3) | 1 (12.5) |
| Hemorrhagic only | 2 (4.1) | 0 (0.0) | 0 (0.0) | 1 (2.3) | 1 (4.5) | 0 (0.0) |
| Both | 17 (34.7) | 10 (43.5) | 10 (38.5) | 16 (37.2) | 8 (36.4) | 6 (75.0) |
| Other combinations | 7 (14.3) | 4 (17.4) | 4 (15.4) | 5 (11.6) | 2 (9.1) | 0 (0.0) |
| Not reported | 13 (26.5) | 6 (26.1) | 7 (26.9) | 13 (30.2) | 5 (22.7) | 1 (12.5) |
| **Other Clinical Information Reported, n (%)** |  |  |  |  |  |  |
| NIH stroke scale | 13 (26.5) | 6 (26.1) | 6 (23.1) | 10 (23.3) | 7 (31.8) | 1 (12.5) |
| Comorbidities | 36 (73.5) | 15 (65.2) | 17 (65.4) | 31 (72.1) | 16 (72.7) | 7 (87.5) |
| Function | 25 (51.0) | 11 (47.8) | 14 (53.8) | 23 (53.5) | 12 (54.5) | 2 (25.0) |
| **Rehabilitation Setting (not mutually exclusive), n (%)** |  |  |  |  |  |  |
| Inpatient Rehabilitation | 22 (44.9) | 9 (39.1) | 10 (38.5) | 19 (44.2) | 7 (31.8) | 4 (50.0) |
| Acute Care Hospital | 17 (34.7) | 6 (26.1) | 5 (19.2) | 14 (32.6) | 8 (36.4) | 2 (25.0) |
| SNF<sup>‡</sup> | 13 (26.5) | 7 (30.4) | 6 (23.1) | 11 (25.6) | 4 (18.2) | 2 (25.0) |
| Outpatient | 12 (24.5) | 7 (30.4) | 8 (30.8) | 12 (27.9) | 6 (27.3) | 2 (25.0) |
| Home Health | 8 (16.3) | 4 (17.4) | 4 (15.4) | 8 (18.6) | 4 (18.2) | 3 (37.5) |
| Long-Term Care Hospital<sup>§</sup> | 1 (2.0) | 0 (0.0) | 0 (0.0) | 1 (2.3) | 0 (0.0) | 0 (0.0) |
| **Provider Type, n (%)** |  |  |  |  |  |  |
| Not specified | 30 (61.2) | 15 (65.1) | 17 (65.4) | 24 (55.8) | 12 (54.4) | 5 (62.5) |
| PT, OT & SLP | 9 (18.4) | 6 (26.1) | 6 (23.1) | 9 (20.9) | 7 (31.8) | 3 (37.5) |
| PT & OT | 8 (16.3) | 1 (4.3) | 2 (7.7) | 8 (18.6) | 2 (9.1) | 0 (0.0) |
| SLP only | 2 (4.1) | 1 (4.3) | 1 (3.8) | 2 (4.7) | 1 (4.5) | 0 (0.0) |
| **Construct Examined (not mutually exclusive), n (%)** |  |  |  |  |  |  |
| Utilization | 35 (71.4) | 14 (60.9) | 17 (65.4) | 32 (74.4) | 15 (68.2) | 4 (50.0) |
| Referral | 13 (26.5) | 7 (30.4) | 6 (23.1) | 10 (23.3) | 5 (22.7) | 3 (37.5) |
| Timeliness of Care | 7 (14.3) | 2 (8.7) | 3 (11.5) | 7 (16.3) | 4 (18.2) | 1 (12.5) |
| **Type of Analysis, n (%)** |  |  |  |  |  |  |
| Bivariate | 12 (24.5) | 2 (8.7) | 4 (15.4) | 9 (20.9) | 1 (4.5) | 0 (0.0) |
| Multivariable | 37 (75.5) | 21 (91.3) | 22 (84.6) | 34 (79.1) | 21 (95.5) | 8 (100.0) |

\*Electronic Health Record
†W = White, B = Black, non-W = non-White
‡Skilled Nursing Facility
§ Long Term Care Hospital
|| PT = Physical Therapy, OT = Occupational Therapy, SLP = Speech-Language Pathology

Twenty-nine studies (59.2%) utilized administrative databases. Electronic health record (EHR) data was the second most common data source (n=15, 30.6%). Most studies (n=36, 73.5%) relied on a single data source, while 13 (26.5%) used more than one. Administrative databases remained the most common data source when stratified by disparities examined, but there were some differences in the distribution of the other data sources. (**Table 1**)

Approximately half of studies used data at the national level (n=24, 49.1%), followed by data at the local and regional levels, respectively. The time periods examined for rehabilitation care delivery ranged from 1990 to 2021. Five studies (10.2%) exclusively covered periods between 1990 and 2000, 23 included periods between 1993 and 2010 (46.9%), 21 (42.9%) included periods between 2003 and 2021, and 5 did not report a time range. (**Supplemental Table S6)**

### Sample Characteristics

#### Sample Size

Ranged from 72 participants to 1.32 million, with a median and interquartile range of 3,889 (643, 15,806) participants (Table 1). When stratified by type of disparity examined, studies reporting on urban/rural disparities had the largest sample sizes.

#### Age

Reporting varied among studies. For those that reported a mean age (n=22), the range was 56-81 years (mean [SD]: 68.4 [5.59]; median [IQR]: 68.1 [65.06, 72.4]). For studies that reported a median age (n=4), the range was 71-72 years. Study populations were, on average, older and in alignment with epidemiologic data on stroke in the US.

#### Sex and Gender

For studies reporting on sex (n=23, 46.9%) and excluding those that included veterans or VA hospitals (N=6, 12.2%), the percentage of female participants ranged from 43.5% to 61.2% (mean [SD]: 52.4% [4.2%]; median [IQR]: 52% [49.7%, 55%]). These percentages are reflective of epidemiologic data that indicate the prevalence of stroke is slightly higher in women, who tend to live longer than men.^40,41^ None of the studies provided information on gender identity.

#### Race/Ethnicity

Most of the studies that reported on race (n=43, 87.9%) used the categories of White/Black (n=19, 38.8%) or White/Black/Other (n=17, 34.7%). Few provided data on other race categories (e.g., Native American, Asian Pacific Islander). For studies reporting the percentage of Black participants (n=40, 81.6%) percentages ranged from 2% to 68% (mean (SD): 23.4% [15.4%]; median [IQR]: 18% [13.2%, 30%]). Of the 20 (40.8%) of studies that assessed Hispanic or Latino ethnicity, percentages ranged from 3% to 39.8% (mean (SD): 10.9% [9.3%]); median [IQR]: 7.6 [6.6%, 9.4%]).

#### Socioeconomic Status

Most studies (n=39, 79.6%) provided data on participants’ insurance, and for most (n=28, 57.1%), this measure could be used as a proxy for SES based on Medicaid enrollment, Medicare-Medicaid dual enrollment, or uninsured/self-pay status. (**Table 1**) Fifteen studies (30.6%) provided information on income. For seven studies (14.3%), income was at the area level (e.g., Census block). Twelve studies (25%) provided information on education, and in most (n=10, 20.4%) the data were at the individual level.

#### Clinical Characteristics

Information varied widely across studies, **(Supplemental Table S7)**, with most reporting on stroke type. Samples including ischemic and hemorrhagic stroke were most common (n=17, 34.7%) (**Table 1**). Studies of participants with hemorrhagic stroke only were least common (n=2, 4.1%). Thirteen (26.5%) reported the NIH stroke scale score, and 36 (76.6%) reported on comorbidities, either a comorbidity index and/or specific comorbidities (e.g., hypertension, cardiovascular disease). Twenty-five (51.0%) reported on patient function. When stratified by disparities examined, fewer studies examining urban/rural disparities controlled for function. The Functional Independence Measure (FIM) was the most common measure of function used (n=8, 16.3%) followed by the Rankin or modified Rankin (n=7, 14.3%).

### Rehabilitation Setting, Provider Type, and Constructs

Most studies (n=32, 65.3%) examined rehabilitation care within a single setting, most commonly IRF (n=22, 44.9%), followed by acute care. (**Figure 3A**, **Table 1**) Only eight (16.3%) examined home-based rehabilitation, and only one examined LTH-based rehabilitation care. Most of the studies that examined rehabilitation care in multiple settings examined acute and post-acute care (i.e., SNF, IRF, HH).

**Figure 3.**
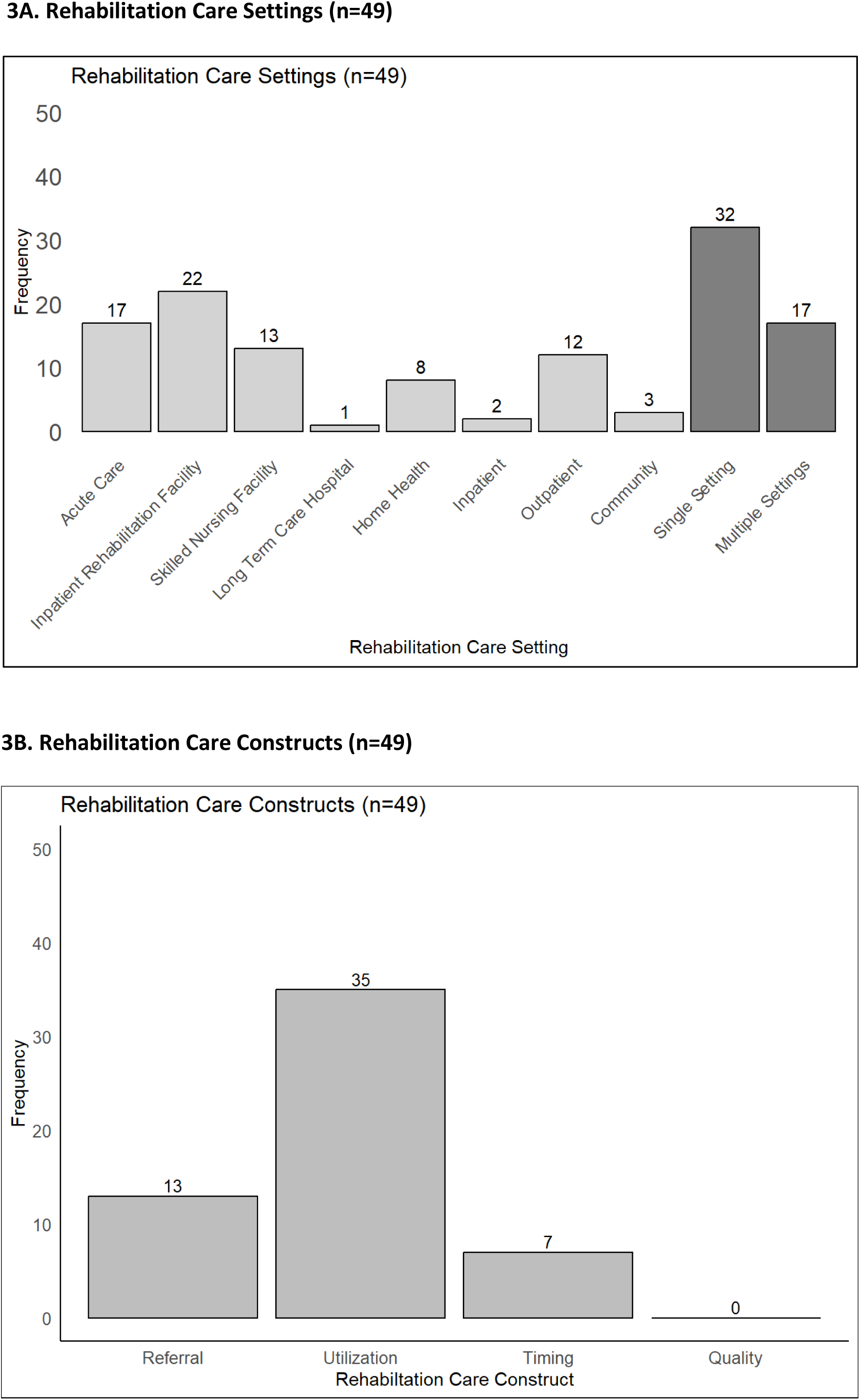
Frequency of Articles in Different Settings and Rehabilitation Care Constructs 3A. Bar chart illustrating the frequency of articles reviewed in various health care settings. **3B.** Bar chart representing the frequency of articles categorized under each rehabilitation care construct.

Most studies (n=30, 61.2%) did not report on rehabilitation provider type. Of those that did, the majority (n=9, 18.4%) reported on the use of PTs, OTs, and SLPs. Eight (16.3%) reported on the use of PTs and OTs only, and 2 focused on SLPs only. (**Table 1**)

Regarding rehabilitation care constructs, most studies (n=35, 71.4%) examined disparities in rehabilitation utilization (i.e., whether or not care was received, number of visits, duration of care). Thirteen (26.5%) examined disparities in referral to rehabilitation, primarily by discharge disposition in claims data, (e.g., discharge to SNF from acute care). Seven (14.3%) examined disparities in the timing of care (e.g., time to start of care relative to stroke onset). (**Table 1**, **Figure 3B**) This trend was similar when stratified by disparities examined. No study examined disparities in rehabilitation care quality. Six (12.2%) examined disparities in two constructs.

### Disparities Examined and Study Findings

Racial disparities were the most common disparity type examined (n=42, 85.7%). (**Figure 4),** with almost half (n=20, 40.8%) operationalizing race and ethnicity as a single variable (e.g., non-Hispanic White, non-Hispanic Black, Hispanic, and other race). Four (8.2%) examined racial and ethnic disparities separately. Twenty-two (45%) examined race only.

**Figure 4.**
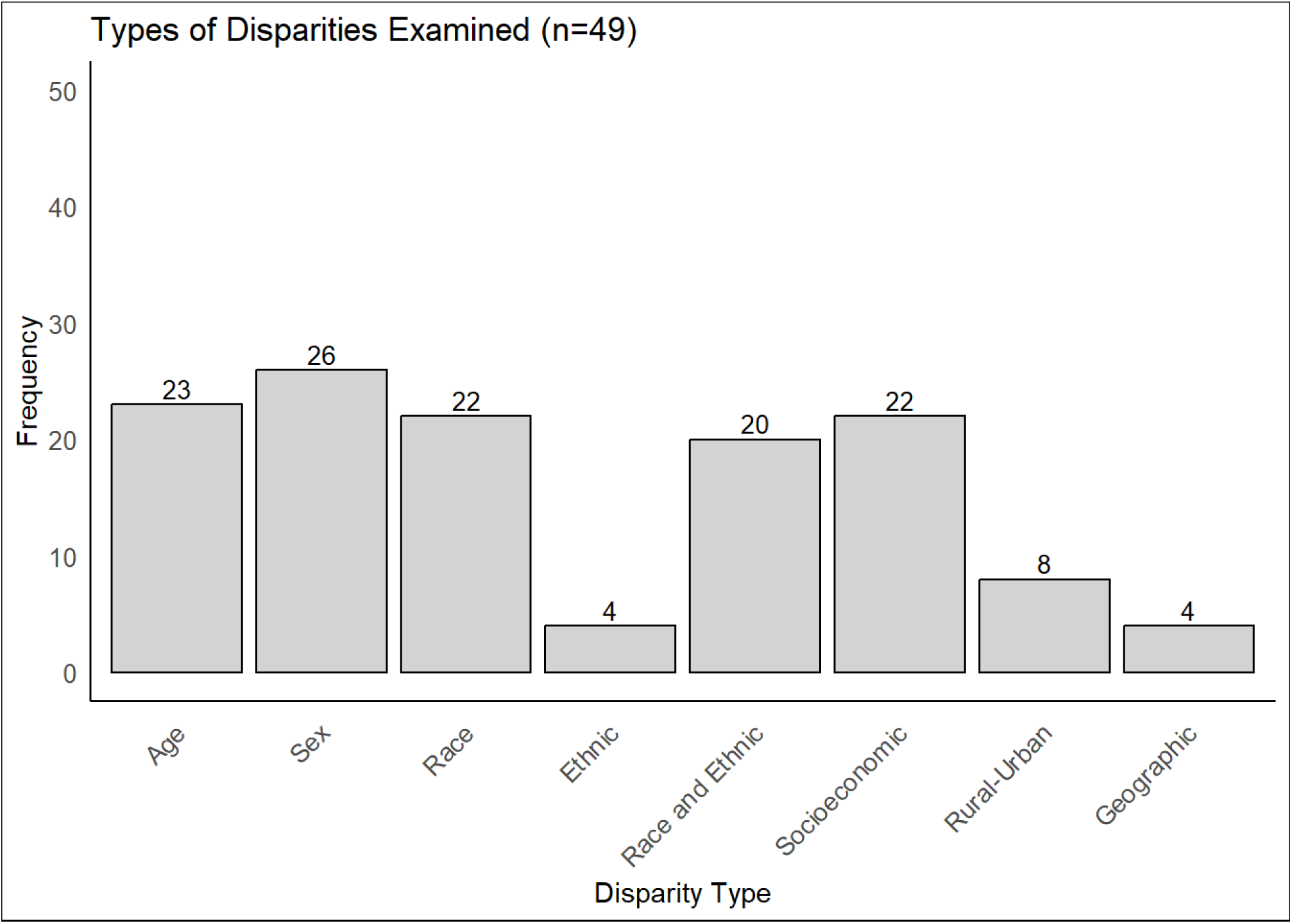
Disparities in Rehabilitation Utilization. This bar chart illustrates the distribution and frequency of reviewed articles in different disparity categories.

Disparities by sex were the next most common type examined (n=26, 53.1% of studies), followed by age and socioeconomic disparities. **(Supplemental Table 8**) Of the 22 studies that examined disparities by SES, the majority (n=15, 68.2%) used insurance status as a proxy for SES, followed by income (n=10, 45.5%), and education (n=6, 27.3%). Only 8 of the 49 studies (16.3%) examined urban/rural disparities, and 4 (8.2%) reported on geographic variation at the state or Census region (tract, block group, or block) level. Most (n=30) examined disparities in more than one category.

Twelve (24.5%) studies reported disparities in rehabilitation care that were unadjusted; 37 (75.5%) used multivariable approaches. (**Table 1**) Most of those with unadjusted results were published before 2011 and examined racial disparities (n=7). These studies reported varied findings with some reporting no disparity, some reporting a disparity for Blacks, and others reporting a disparity for Whites (**Supplemental Table S8**).

A total of 63 rehabilitation care outcomes were examined across the 37 studies that utilized multivariable analyses to examine disparities. These studies controlled, to at least some degree, for patient characteristics that may influence therapy use. Most of the outcomes examined were related to use of care (68%), referral (22%), and timing (10%).

**Figure 5A** summarizes findings from the studies that used multivariable analyses. We defined a disparity as not receiving care, receiving less care (i.e., fewer visits, shorter duration of care, or less intensive care [e.g., referral to SNF vs IRF]), or receiving care in a less timely manner. As the figure illustrates, there was a large amount of variability in whether a disparity was identified and the direction of the disparity. For example, of the 31 multivariable analyses that evaluated age disparities, 9 (32%) found no disparity, 12 (39%) reported older people received less rehabilitation, and 10 (29%) reported younger people received less rehabilitation. Studies that identified a sex disparity more often found that females received less rehabilitation; however, almost half found no difference by sex. For many studies, there were no racial or ethnic disparities or there were racial/ethnic disparities with Whites or Non-Hispanics receiving less care. The most consistent findings indicate disparities in rehabilitation care based on lower SES and rural residence. Furthermore, of the seven analyses that examined the association between geography and rehabilitation care, all reported variation by geographic region, such as state or Census region (data not included in figure).

**Figure 5:**
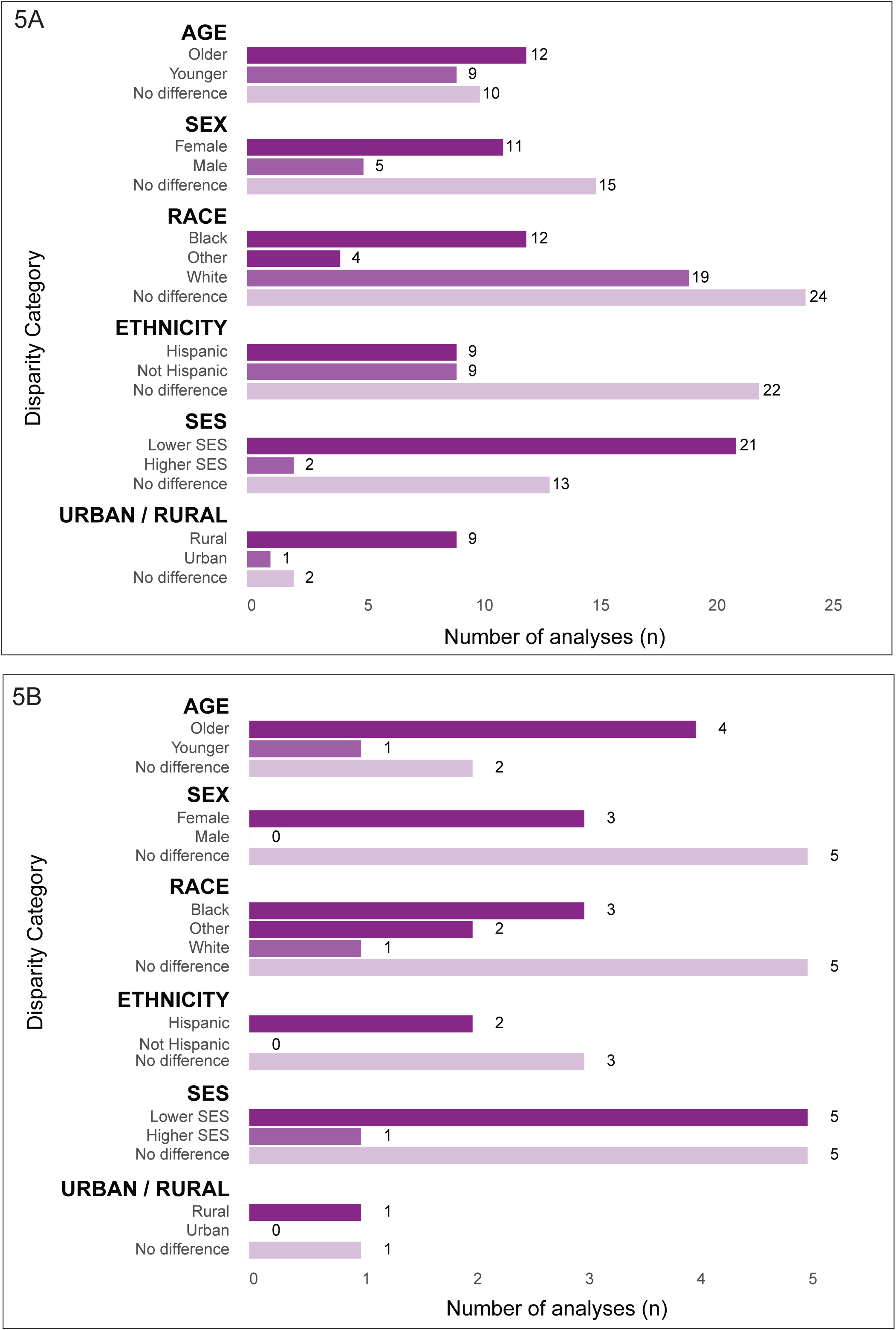
Clustered Bar Charts Showing Disparity Findings in Different Categories. 5A. Clustered bar chart representing the number of articles that found a disparity in various categories, based on studies that employed multivariable analyses. 5B. Clustered bar chart showing the number of articles identifying a disparity in different categories, specifically from studies that controlled for functional status, highlighting more consistent findings.

Subgroup analyses by rehabilitation care construct (i.e., referral, use, timing) did not reveal any trends that were different than what was seen in the overall analysis. When examining disparity findings based on setting, some trends became apparent. For the analyses of sex-based disparities in outpatient rehabilitation care, the majority indicated a disparity for females; for racial disparities in acute care, a majority indicated no disparities; and for use of IRF care (vs SNF care), a majority indicated a disparity for older adults and females. (**Supplemental Table 8**)

Limiting the analyses to those that controlled for function (N=15), some of the differences seen for groups that are not considered disparity populations (e.g., younger, male, White, etc.) were attenuated (e.g., only one of the seven analyses that examined age disparities found that younger patients were less likely to receive care). Similar trends were seen by sex, race, and ethnicity.

## DISCUSSION

The objectives of this scoping review were to summarize current evidence on disparities in rehabilitation care following stroke, identify gaps in the literature, and make recommendations for future research. Findings from the studies we reviewed varied, with the most consistent findings indicating that individuals with lower SES and those living in rural areas (vs urban) are less likely to receive rehabilitation care following stroke. Studies that examined rehabilitation care delivery by Census region or state also consistently reported variation, suggesting potential disparities in care based on one residence. Findings were mixed for studies examining disparities in age, sex, race, and ethnicity, with some studies reporting a disparity in one direction, others reporting no disparities, and still others reporting disparities in the opposite direction. When we limited findings to analyses that controlled for functional status, findings across studies were more consistent (**Figure 5B**). Most studies examining disparities in age and race indicated that older individuals and non-White individuals were less likely to receive rehabilitation care, while most studies examining disparities in sex and ethnicity indicated no disparities in rehabilitation care.

The varied findings of our scoping review underscore the importance of considering the methodologic quality of studies when summarizing evidence. When examining disparities in rehabilitation care it is important to control for the need for therapy through measures such as functional status and ability to perform activities of daily living. While assessing study quality was not an objective of this review, our findings indicate the need to meta-analyze results and assess the risk of bias.

Some of the variation in results is also likely due to variation in the characteristics of the samples examined (e.g., by age, stroke type, time since stroke); the numerous ways in which disparities in rehabilitation care following stroke can be examined based on the data source and the setting(s) and provider types examined. There are also different ways to examine rehabilitation care delivery (e.g., receipt of care, frequency, intensity, duration, quality). Such complexity makes conducting research on this topic and summarizing the evidence challenging.

Based on our assessment of the evidence, many gaps remain in our understanding of disparities in rehabilitation care. While there has been more interest in moving beyond mere identification of healthcare disparities and towards efforts to minimize or eliminate these disparities, identification of the type and extent of disparities is a necessary first step. From a methodologic perspective, we have several recommendations:

- **Data Source**. The data source should be representative of the populations of interest and as current as possible considering the rapidly changing healthcare landscape, particularly in the area of post-acute care. Using billing data from a healthcare system or payer (e.g., administrative Medicare claims data) provides comprehensive, valid, and representative data on healthcare use. These data alone, however, do not provide details on the content of the rehabilitation care (e.g., the specific treatments used to address balance problems) or the individual’s functional status, which is strong measure of “need” for therapy. These data may also have other limitations such as missingness and measurement errors in race/ethnicity (e.g., Medicare claims data include Hispanic ethnicity as a race category).
- **Supplemental Data**. To conduct meaningful analyses, billing or administrative data should be supplemented with other data from the EHR and/or other sources such as Census tract or registry data to provide additional information on demographics, functional status, content of rehabilitation, family support, patient preferences, and social determinants of health. These data are often more challenging to obtain but are critical for fully understanding the reasons behind the observed disparities.
- **Health Disparity Populations.** Sociodemographic categories should be clearly and appropriately defined at the individual level (vs an area level measure) following current conventions and/or recommendations and inclusive. We identified several studies that treated Hispanic/Latino ethnicity as a race category. Few studies specifically examined ethnic disparities in rehabilitation use. No study addressed gender disparities. The way SES was captured also varied across studies with the majority using insurance as a proxy for SES and some measuring SES at the area level and not the individual level. Finally, few studies examined urban/rural disparities in rehabilitation care.
- **Rehabilitation Care Settings and Provider Types.** We found that most of the studies examined disparities in the inpatient settings of the acute care hospital, IRF, and SNF. Subgroup analyses also suggested fewer racial disparities in these settings. Based on those findings and the current emphasis on shifting care to the community as quickly and as much as possible, we recommend that future studies focus on understanding disparities in rehabilitation care in the home and outpatient settings and in the transition from the institution to the community (e.g., disparities in the time from discharge home to the start of care). More studies that specifically look at access to each provider type (i.e., PT, OT, SLP) are also needed to understand the extent to which disparities may vary by provider type and how this may impact outcomes.
- **Rehabilitation Care Constructs.** Most studies we reviewed examined disparities in rehabilitation utilization and operationalized rehabilitation use as a dichotomous variable (received or did not receive). While this information, as well as information on disparities in referral to rehabilitation, are important, more studies are needed that look at disparities in the timing of care; the frequency, duration, and intensity of care; and the types of interventions used. Disparities in the quality of care, which is a more challenging construct to operationalize, is ultimately the most important construct to examine. More studies should attempt to operationalize and examine disparities in the quality of care. For example, the IOM defines high quality care as care that is safe, effective, patient-centered, timely, efficient, and equitable.^42^ One possible way to operationalize high quality care is care that is timely (e.g., received within 7 days of discharge home) and effective (e.g., interventions used are evidence-based).
- **Context.** Most of the studies used national-level data to examine disparities in rehabilitation care. Findings from such studies are particularly useful for informing policy at the national level but are less actionable from a local context. We recommend more studies using local or regional health system data, where disparities can be identified and serve as a launching point for understanding the multi-level factors that may contribute to these disparities and intervening on these factors. These may include individual-level factors related to the patient, the healthcare provider, and healthcare administrators; organizational-level factors such as infrastructure and culture; and community-level factors such as supply of healthcare providers and availability of public transportation. Understanding factors that contribute to disparities at the local level will provide the most useful information for developing interventions to begin to minimize or eliminate these disparities.

Additional research that addresses these gaps is essential and supports the WHO’s Rehabilitation 2030 Initiative, which identifies rehabilitation as an essential health service. This initiative sets forth the goal of “building comprehensive rehabilitation service delivery models that progressively achieve equitable access to quality services.”^43^ This goal is particularly relevant for patients with stroke, as the incidence is expected to increase as the population ages. Stroke also disproportionately affects racial minorities, women, and individuals of lower SES.^44–46^ In the US, Black adults are 50% more likely to have a stroke than their White counterparts. Furthermore, Black Americans are 70% more likely to die from a stroke than their non-Hispanic White counterparts, and those that do survive are more likely to have residual physical disability. The lifetime risk of stroke is also higher among American women (20%–21%) than men (14%– 17%).^40,47^ Further, the age of onset of stroke is 4-6 years older among women than men, leaving them more likely to have a higher degree of disability post-stroke and more likely to be living alone.^47–50^ Those with lower SES have higher stroke incidence and mortality^51^ as well as higher stroke severity^52^ and post-stroke disability.^53^

Practice guidelines for stroke rehabilitation are well established and recommend that stroke survivors receive rehabilitation at an intensity commensurate with anticipated benefit and tolerance.^54^ However, access to rehabilitation services remains a major barrier. Availability of post-acute care settings (especially in underserved areas), prospective payment variability, regulatory practices, the pressure to discharge patients rapidly from acute care, and sociodemographic characteristics of patients all may influence if and where rehabilitation services are provided.^55^ While the extent of racial, ethnic, sex, and gender identity disparities in rehabilitation following stroke was not clear based on this review, the consistency in findings regarding SES and rural status (which likely serves as a proxy for provider availability) highlight potential areas to target in future work. Indeed, the AHA/ASA policy statement on stroke systems of care underscores the multiple barriers patients face in receiving rehabilitation care across the continuum. One of their recommendations for stroke systems of care is that Federal and other governmental institutions enact policies to “facilitate access to secondary prevention and rehabilitation and recovery resources after stroke.” A second recommendation states that “A stroke system should periodically assess its level of available rehabilitation services and community resources.”

### Limitations

There are several limitations to this review. Due to the nature of a scoping review, we adopted broad inclusion criteria, including studies that did not state an objective of examining disparities in rehabilitation care. While this allowed for a comprehensive view of the current literature, it may have introduced peripheral findings and may be one explanation for analyses that lacked appropriate methodologies for examining disparities in care. Second, we limited our search to studies conducted in the US given the strong social, geographical, and political influences that shape disparities in healthcare. This may have limited our discovery of additional methodologies/issues to examine disparities in rehabilitation care. Third, we did not perform metanalyses or assess studies for rigor as this is not within the purview of a scoping review. Fourth, we examined data across multiple constructs of healthcare including different settings, different constructs of care (i.e., referral, timing, utilization, quality), and different disparity populations. While this provided a descriptive overview of the state of the current literature on this topic, it was complex to summarize and synthesize the findings from these articles and systematically identify all of the gaps in the literature. Summarizing the literature via a systematic review and meta-analysis would likely bring more clarity to this topic.

### Conclusion

We conducted a scoping review and identified 49 articles that examined disparities in rehabilitation care following stroke. Racial, sex, and age disparities were the most frequently examined. We found consistent reports of less rehabilitation care use by individuals of lower SES and living in rural areas. While findings on racial, sex, and age disparities varied, when limiting our analysis to studies that used multivariable approaches to control for the “need” for therapy, most studies suggested disparities in rehabilitation care for non-White individuals and older individuals and no disparities by sex or ethnicity. We identified many gaps in our understanding of disparities in rehabilitation care following stroke that can be addressed in future studies.

## Data Availability

The data that support the findings of this study are available on request from the corresponding author.

## ACKNOWLEDGEMENTS

None

